# Utilizing pharmacy refill data to predict loss to follow-up among people living with HIV in Manyara region of Tanzania

**DOI:** 10.64898/2026.02.24.26347034

**Authors:** Michael B Kalulo, Raphael Z. Sangeda, James Mwakyomo, George R. Sangeda, Veryeh Sambu, Prosper Njau

## Abstract

**Background:** Achieving optimal adherence and retention in HIV care is essential for sustaining viral suppression. Pharmacy refill records offer an objective approach to assessing adherence in settings where routine viral load testing is limited. This study evaluated pharmacy refill adherence, loss to follow-up (LTFU), and their predictors among people living with HIV (PLHIV) in the Manyara region of Tanzania.

**Methods:** We conducted a retrospective cohort analysis of 22,650 PLHIV across five districts using the National CTC-2 database. LTFU was defined as no clinic visit for 180 days or more. We also analyzed a cross-sectional final status field updated by health trackers to distinguish research-defined LTFU from confirmed clinical outcomes. Predictors were evaluated using multivariable logistic regression, and spatial mapping identified geographic disparities.

**Results:** The mean pharmacy refill adherence was 84.1%, with 57.9% achieving good adherence (>=85%). In the longitudinal analysis, 32.8% of patients met the research definition for LTFU (>=180-day absence) at some point during the study period. Cumulative LTFU was significantly higher in earlier initiation cohorts (2017-2019) compared to the 2021 cohort (aOR 1.89; 95% CI 1.76-2.02). However, cross-sectional system records, which health trackers update, showed that only 2.9% remained truly lost to care; 65.3% were active at their original clinic, 23.1% had eventually transferred to other facilities, and 6.7% were deceased. In multivariable regression, poor pharmacy adherence was the strongest behavioral predictor of disengagement (aOR 2.04; 95% CI 1.77-2.35). Significant geographic variation was observed, with residence in Simanjiro independently associated with the highest odds of LTFU (aOR 3.60; 95% CI 2.67-4.85). Spatial mapping confirmed a clustering of poor outcomes in districts characterized by nomadic pastoralist livelihoods.

**Conclusion:** Pharmacy refill adherence is a potent predictor of disengagement and a practical early-warning indicator. The high rate of silent transfers and district-level disparities, particularly in nomadic hotspots, highlight the need for a national unique patient identifier and mobility-friendly retention strategies. Integrating automated refill alerts into the 90-day tracking window is essential to achieve 95-95-95 targets.

## Introduction

Globally, HIV remains a major public health concern, with an estimated 40.8 million people living with HIV (PLHIV) in 2024 [1]. Eastern and Southern Africa continue to experience the highest burden of HIV, although notable progress has been made through antiretroviral therapy (ART) scale-up and differentiated service delivery (DSD) [2,3]. Tanzania has experienced a rapid expansion of ART coverage and a major transition toward DTG-based regimens in recent years, as evidenced by national dispensing data showing more than a doubling of ART consumption from 2017 to 2021[4]. In Tanzania, approximately 1.7 million people live with HIV, with substantial geographic heterogeneity in prevalence. According to the Tanzania HIV Impact Survey *THIS 2022-2023* [5], Manyara is one of the lowest-prevalence regions, with only 1.5% of adults aged 15-49 years infected, which is much lower than high-burden regions such as Njombe (8.4%), Iringa (6.3%) and Mbeya (6.1%) [5].

Global HIV control efforts are anchored in the UNAIDS 95-95-95 targets for 2030, which aim for 95% of all PLHIV to know their status, 95% of those diagnosed to be on ART, and 95% of those on treatment to achieve viral suppression [1,6]. Achieving the final target depends heavily on sustained ART adherence and retention. High levels of adherence, typically ≥85-95%, are required to maintain viral suppression, prevent disease progression, and minimize onward transmission [7,8].

Several approaches exist for measuring adherence, including pill counts, self-reports, appointment adherence, and electronic monitoring. However, these methods vary in terms of reliability and feasibility, particularly in resource-limited settings. Pharmacy refill adherence has emerged as an objective, low-cost, and scalable indicator of medication-taking behaviors. Evidence from Tanzania demonstrates that pharmacy refill-based adherence more accurately predicts virological outcomes compared with self-reported adherence [9,10]. Similar findings have been reported in Brazil [11].

Loss to follow-up (LTFU) remains a major challenge for sustained ART effectiveness. In sub-Saharan Africa, 20-25% of PLHIV become LTFU within 12 months of ART initiation, far exceeding the WHO targets [12,13]. In Tanzania, studies have reported LTFU levels ranging from 20% to over 30% depending on the region and patient population [13,14]. Factors contributing to LTFU include younger age, stigma, socio-economic hardship, mobility, poor adherence, and limited psychosocial support [15,16].

Although pharmacy refill data have been used to monitor adherence in urban Tanzanian cohorts [9,17], there is limited evidence of their utility in predicting LTFU in low-prevalence regions, such as Manyara. Moreover, district-level spatial heterogeneity in adherence and retention has not been mapped or analyzed in this region. Understanding such variations is essential for tailoring DSD models, strengthening retention strategies, and optimizing resource allocation.

Therefore, this study aimed to evaluate pharmacy refill adherence and LTFU among PLHIV receiving ART in the Manyara region and identify demographic, geographic, and temporal predictors of LTFU using logistic regression. We also examined whether refill-based adherence could reliably predict disengagement from care and used geospatial mapping to visualize district-level patterns of adherence and retention. Together, these analyses provide insights into the individual and spatial determinants of LTFU and the potential utility of pharmacy refill data within routine HIV program monitoring.

## Methods

### Study design and setting

This study was a secondary analysis of routinely collected HIV program data from the National AIDS and STI Control Programme (NASHCoP) in Tanzania. The analytic period spanned January 2017 to December 2021, representing the most complete interval with reliable pharmacy refill, follow-up, and viral load information. The analysis focused on the Manyara region in northern Tanzania and included all care and treatment centers (CTCs) in the districts of Babati, Hanang, Kiteto, Mbulu, and Simanjiro.

### Study population and eligibility criteria

The study population comprised all children, adolescents and adults living with HIV who were enrolled in care and receiving ART in Manyara during the study period. A complete census of all eligible individuals was conducted, yielding 22,650 participants in the study. Inclusion required at least one recorded ART refill and valid patient identification. Viral load analyses were restricted to individuals with at least one documented viral load result; however, viral load availability was not required for inclusion in the adherence or LTFU analyses.

### Data sources and variables

Data were extracted from the national CTC-2 database maintained by NASHCoP, which contains longitudinal demographic, clinical, laboratory, and pharmacy dispensing information for PLHIV in Tanzania. The extracted variables included age, gender, marital status, district of residence, date of ART initiation, regimen history, CD4 count, viral load results, scheduled appointment dates, number of tablets dispensed, days of medication supplied, and programmatic outcomes at the last recorded visit (active, transferred out, died, lost to follow-up, or opted out). Pharmacy dispensing records were used to reconstruct medication possession patterns.

### Computation of pharmacy refill adherence

Pharmacy refill adherence was quantified using a visit-based, time-dependent approach that assessed medication possession across consecutive refill intervals. For each individual, the number of days late relative to the scheduled pharmacy refill dates was calculated at every visit and cumulatively summed over the entire follow-up period. Adherence was expressed as the proportion of time during which medication was available using the following formula:

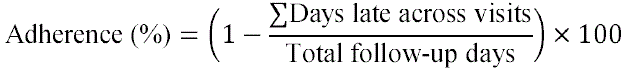

This allows adherence to be computed at the visit level and aggregated over any defined period (e.g., annually or across the complete follow-up). Adherence values were expressed on a 0-100% scale and rounded to the nearest whole percentage point.

In accordance with national HIV program guidelines and consistent with prior validation studies demonstrating the predictive performance of pharmacy refill-based adherence for treatment outcomes and LFTU in resource-limited settings [9,10], adherence of ≥85% was classified as good and adherence of <85% as poor.

### Definition of Loss to follow-up

LTFU was defined according to the standards established by the NASHCoP. While recent national guidelines have prioritized a 90-day threshold for early programmatic intervention, this study utilized a more conservative research definition: patients were classified as LTFU if no clinic visit was recorded for 180 days or more after their last scheduled appointment. This window was selected to ensure that patients were not simply late for an appointment but had definitively disengaged from care at their primary facility.

In the multivariable logistic regression models, patients documented as having died, been formally transferred to another health facility, or voluntarily opted out of care were categorized separately and excluded from the LTFU outcome. Additionally, we reviewed the cross-sectional final status field, which includes updates from active health tracking and tracing efforts, to distinguish between those meeting the 180-day research definition of loss and those with confirmed clinical outcomes, such as silent transfers or mortality.

### Data management and statistical analysis

Data cleaning and preprocessing were performed in R for temporal structuring, computation of derived variables, geospatial analysis, and descriptive and inferential statistics. The preprocessing steps included chronological ordering of clinic visits, reconstruction of individual follow-up histories, calculation of pharmacy refill adherence, assignment of LTFU status, and linkage of viral load results to the corresponding patient records.

Descriptive statistics (means, standard deviations, frequencies, and proportions) were used to summarize the cohort’s demographic and clinical characteristics. The spatial distributions of adherence and LTFU across districts in the Manyara region were analyzed and visualized using the sf, ggplot2, ggspatial, and viridis packages in R, allowing for district-level geospatial comparisons.

Associations between categorical variables were examined using Pearson’s chi-square test. Logistic regression models were used to identify the predictors of LTFU. Univariate logistic regression was first performed for each potential predictor; variables with p < 0.20 in the univariate stage were retained for multivariable analysis to avoid premature exclusion of variables with confounding effects. Adjusted odds ratios (aORs) with 95% confidence intervals were reported for the multivariable models. Statistical significance was set at p < 0.05.

### Ethical considerations

This study used de-identified routine program data from the NASHCoP. Ethical approval for the secondary analysis of CTC-2 data was obtained from the Muhimbili University of Health and Allied Sciences (MUHAS) Institutional Review Board under protocol number DA.282/298/01L/923 issued in February 2024.

## Results

### Study population

A total of 22,650 PLHIV receiving ART in the Manyara region were included in the study. The cohort was predominantly female (68.7%) and aged 29-69 years (72.4%). District representation was highest in Babati (32.0%), followed by Kiteto (23.4%) and Simanjiro (23.0%). The mean baseline age was 37.5 years, increasing to 39.5 years at the end of follow-up. The mean follow-up duration was 844 days (Table 1).

**Table 1:** Demographic and clinical characteristics of PLHIV in Manyara region (N = 22,650)

| Characteristic | Category | N (%) |
| --- | --- | --- |
| **Gender** | Male | 7,087 (31.3%) |
|  | Female | 15,563 (68.7%) |
| **Age Group** | 0-10 | 1,007 (4.4%) |
|  | 11-18 | 848 (3.7%) |
|  | 19-28 | 3,993 (17.6%) |
|  | 29-69 | 16,402 (72.4%) |
|  | ≥70 | 400 (1.8%) |
| **District** | Babati | 7,251 (32.0%) |
|  | Kiteto | 5,309 (23.4%) |
|  | Simanjiro | 5,219 (23.0%) |
|  | Mbulu | 3,008 (13.3%) |
|  | Hanang | 1,863 (8.2%) |
| **Marital Status** | Married | 11,209 (49.5%) |
|  | Single | 4,230 (18.7%) |
|  | Divorced | 1,783 (7.9%) |
|  | Widowed | 1,158 (5.1%) |
|  | Child | 484 (2.1%) |
|  | Cohabiting | 237 (1.0%) |
|  | Not specified | 3,548 (15.7%) |
| **Continuous Variables** | Mean age (years) | 37.5 (SD 14.6) |
|  | Mean age at end (years) | 39.5 (SD 14.8) |
|  | Follow-up duration (days) | 844 (SD 652) |

### Pharmacy refill adherence

The mean pharmacy refill adherence was 84.1% (SD 18.3). Overall, 57.9% of the participants achieved good adherence (≥85%), whereas 42.1% had poor adherence (Table 2).

**Table 2:**
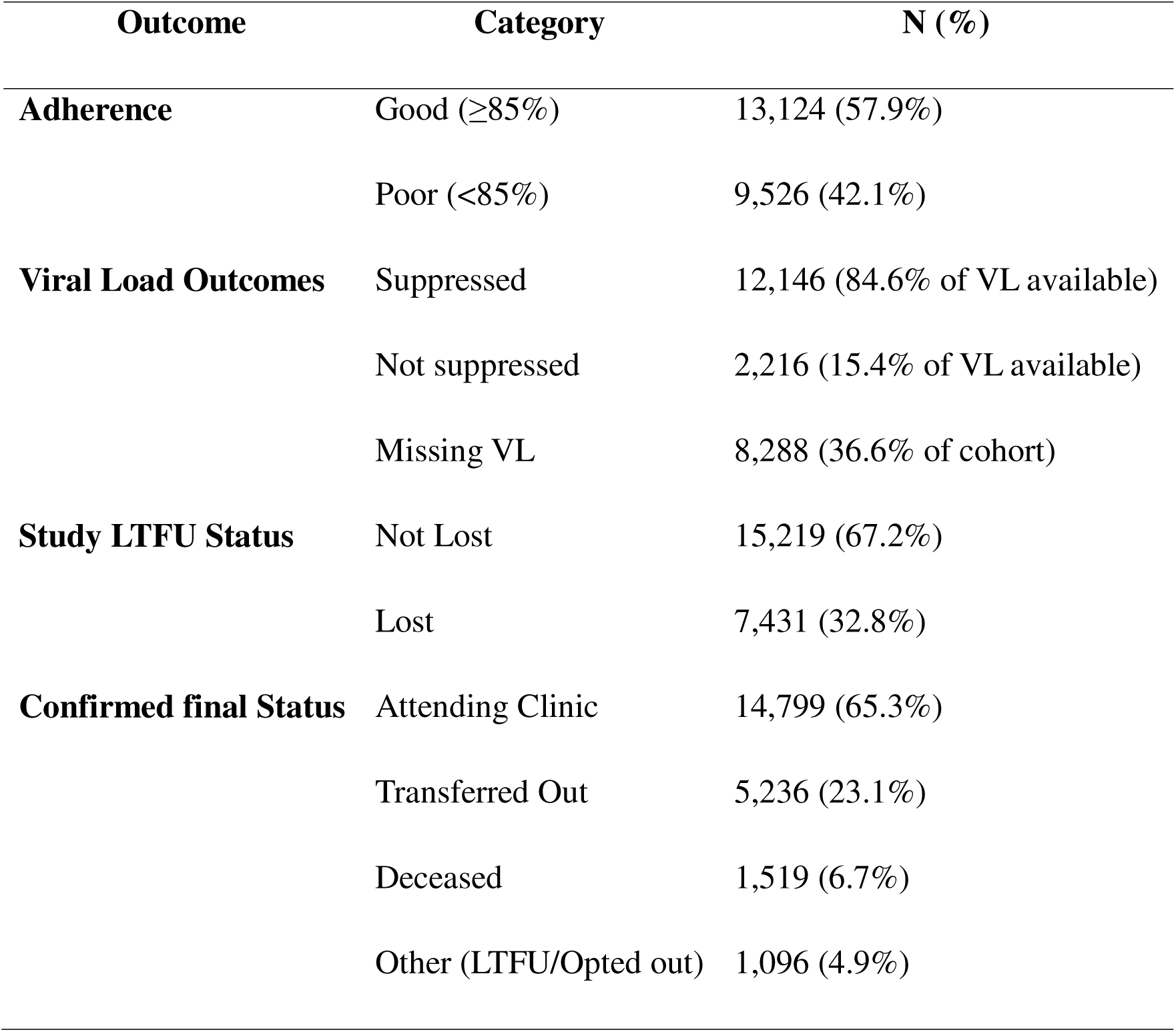
Adherence, viral load outcomes, and Loss to follow-up.

| **Outcome** | **Category** | **N (%)** |
| --- | --- | --- |
| **Adherence** | Good ($\geq 85\%$) | 13,124 (57.9%) |
| | Poor ($< 85\%$) | 9,526 (42.1%) |
| **Viral Load Outcomes** | Suppressed | 12,146 (84.6% of VL available) |
|  | Not suppressed | 2,216 (15.4% of VL available) |
|  | Missing VL | 8,288 (36.6% of cohort) |
| **Study LTFU Status** | Not Lost | 15,219 (67.2%) |
|  | Lost | 7,431 (32.8%) |
| **Confirmed final Status** | Attending Clinic | 14,799 (65.3%) |
|  | Transferred Out | 5,236 (23.1%) |
|  | Deceased | 1,519 (6.7%) |
|  | Other (LTFU/Opted out) | 1,096 (4.9%) |

### Loss To Follow-Up for Six Months

Based on the research definition of no clinic contact for 180 days or more, 32.8% (n = 7,431) of the cohort were classified as LTFU. The remaining 67.2% (n = 15,219) were considered retained in care at their primary facility during the study period (Table 2).

### Confirmed Final Status

To provide a comprehensive clinical outcome, we evaluated the final status field, which incorporates updates from health trackers and tracing teams. These cross-sectional records confirmed that 65.3% (n = 14,799) of the total cohort were active at their original clinic. A significant proportion of the remaining patients (23.1%, n = 5,236) were documented as having transferred care to another facility, while 6.7% (n = 1,519) were confirmed deceased. Following these system-wide updates, only 2.9% (n = 656) remained officially categorized as lost in the system after tracing efforts (Table 2).

### Spatial distribution of LTFU and adherence

Substantial geographic heterogeneity was observed (Table 3, with the Simanjiro district recording the highest LTFU (46.2%), followed by Kiteto (31.1%) and Hanang (30.5%). In contrast, the Babati district demonstrated the lowest LTFU (26.5%). The distribution of pharmacy refill adherence mirrored this pattern: Simanjiro again had the lowest mean adherence (78.35%), followed by Hanang (87.51%), Mbulu, and Babati (Figure 1), respectively.

**Figure 1:**
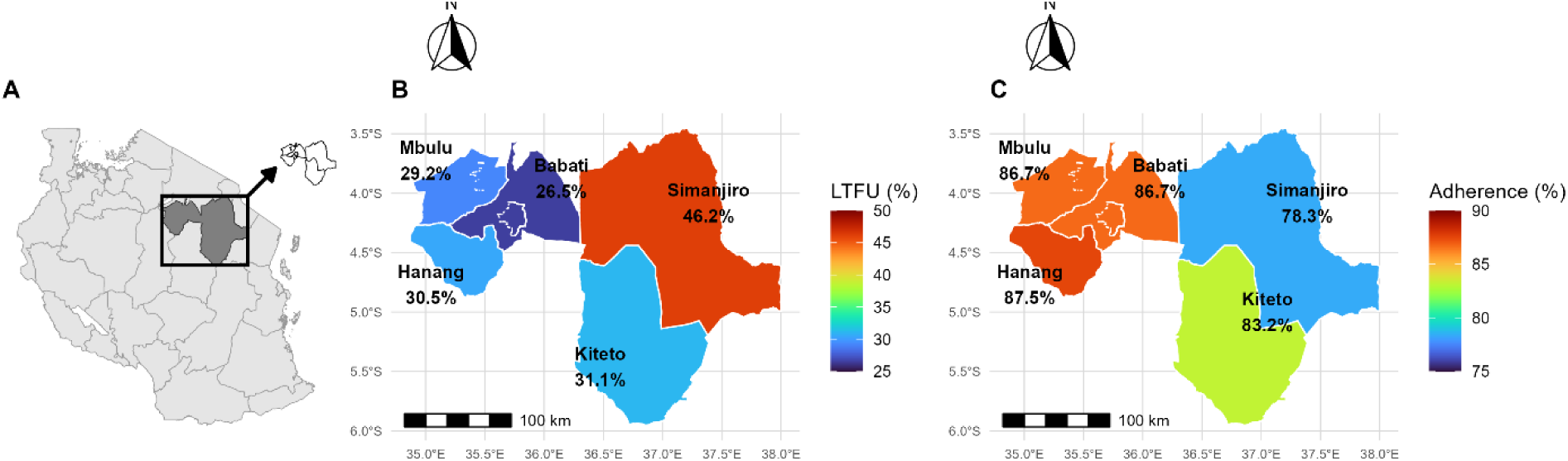
Spatial distribution of loss to follow-up and pharmacy refill adherence in the Manyara Region of Tanzania. Panel A: Map of Tanzania showing the location of the Manyara region (highlighted), with an inset displaying the five study districts: Babati, Kiteto, Simanjiro, Mbulu, and Hanang. Panel B: District-level Loss to follow-up (LTFU) percentages; Panel C: District-level mean pharmacy refill adherence.

**Table 3:** District-level mean adherence and loss to follow-up in Manyara region.

| **District** | **Mean adherence (%)** | **LTFU (%)** |
| --- | --- | --- |
| Babati | 86.70 | 26.5 |
| Kiteto | 83.19 | 31.1 |
| Simanjiro | 78.35 | 46.2 |
| Mbulu | 86.68 | 29.2 |
| Hanang | 87.51 | 30.5 |

### Viral load outcomes

Viral load (VL) results were available for 14,362 patients in this study. Among those with VL results, 84.6% achieved virological suppression, and 15.4% experienced virological failure (Table 2). VL data were missing for 36.6% of the cohort due to programmatic exclusion.

### Temporal trends in loss to follow-up by ART initiation cohort (Figure 2)

Temporally, LTFU peaked among those who initiated ART in 2018 (45.7%) and 2019 (43.6%), with a significant reduction in the 2021 cohort (6.8%), reflecting both shorter follow-up and improved retention strategies (Figure 2).

**Figure 2:**
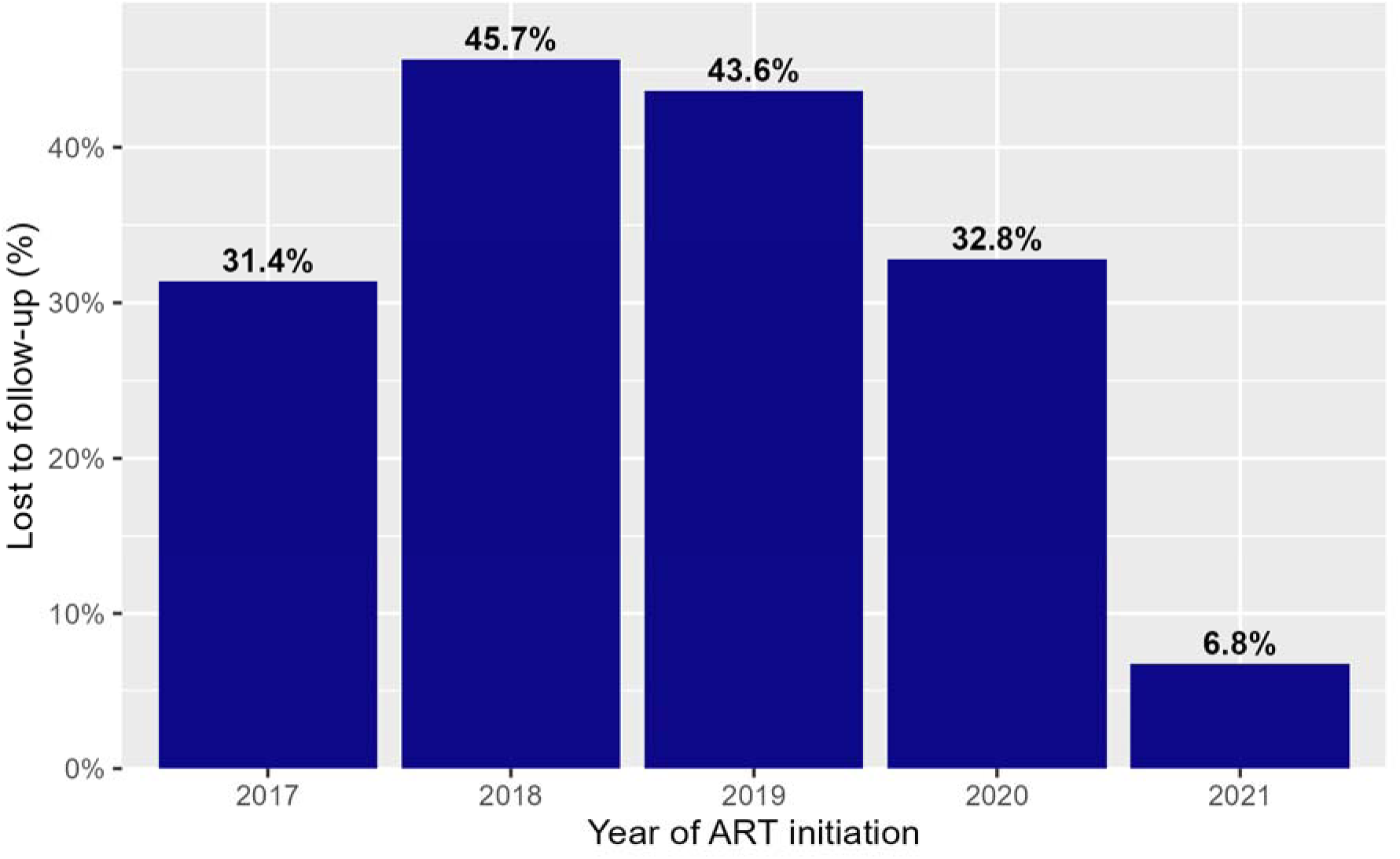
Proportion of people living with HIV lost to follow-up by year of ART initiation in the Manyara region (2017-2021). LTFU was highest among individuals initiating ART in 2018 and 2019 (45.7% and 43.6%, respectively) and lowest in the 2021 cohort (6.8%).

LTFU peaked among individuals who initiated ART in 2018 (45.7%) and 2019 (43.6%), with moderate levels observed in the 2017 and 2020 cohorts. A substantial improvement was observed among those who initiated ART in 2021, with LTFU dropping to 6.8%, likely reflecting the shorter follow-up duration and improvements in retention strategies implemented in recent years (Figure 2).

### Predictors of Loss to Follow-Up

In a univariate analysis (Table 4), poor adherence (OR 3.49), residence in Simanjiro (OR 1.95), and an earlier year of initiation (2018; OR 11.60) were strongly associated with LTFU.

**Table 4:** Univariate Predictors of Loss to Follow-Up (LTFU) Among PLHIV in the Manyara Region.

| Predictor | Category / Unit | OR | 95% CI | p-value |
| --- | --- | --- | --- | --- |
| Age | Per year increase | 0.99 | 0.99 - 1.00 | <0.001 |
| Gender | Male vs. Female | 1.04 | 0.98 - 1.11 | 0.180 |
| District | Babati | Reference | - | - |
|  | Kiteto | 0.82 | 0.73 - 0.92 | 0.001 |
|  | Simanjiro | 1.95 | 1.75 - 2.19 | <0.001 |
|  | Mbulu | 0.94 | 0.83 - 1.06 | 0.315 |
|  | Hanang | 1.22 | 1.06 - 1.39 | 0.001 |
| Adherence | Poor (<85%) vs. Good | 3.49 | 3.29 - 3.70 | <0.001 |
| ART Initiation | 2021 | Reference | - | - |
|  | 2017 | 6.32 | 5.38 - 7.42 | <0.001 |
|  | 2018 | 11.60 | 9.43 - 14.26 | <0.001 |
|  | 2019 | 10.68 | 8.67 - 13.14 | <0.001 |
|  | 2020 | 6.74 | 5.47 - 8.30 | <0.001 |
| Follow-up | Per day increase | 0.998 | 0.997 - 0.999 | <0.001 |

In the multivariable model (Table 5), poor pharmacy refill adherence remained a significant behavioral predictor (aOR 2.04; 95% CI 1.77-2.35). Geographic location emerged as the strongest independent risk factor, with patients in Simanjiro having more than triple the odds of LTFU compared with those in Babati (aOR 3.60; 95% CI 2.67-4.85). Furthermore, patients who initiated ART during 2017-2019 had nearly twice the odds of LTFU compared to the 2020-2021 cohort (aOR 1.89; 95% CI 1.76-2.02). In this adjusted model, male patients showed slightly lower odds of LTFU than females (aOR 0.88; 95% CI 0.83-0.94).

**Table 5:**
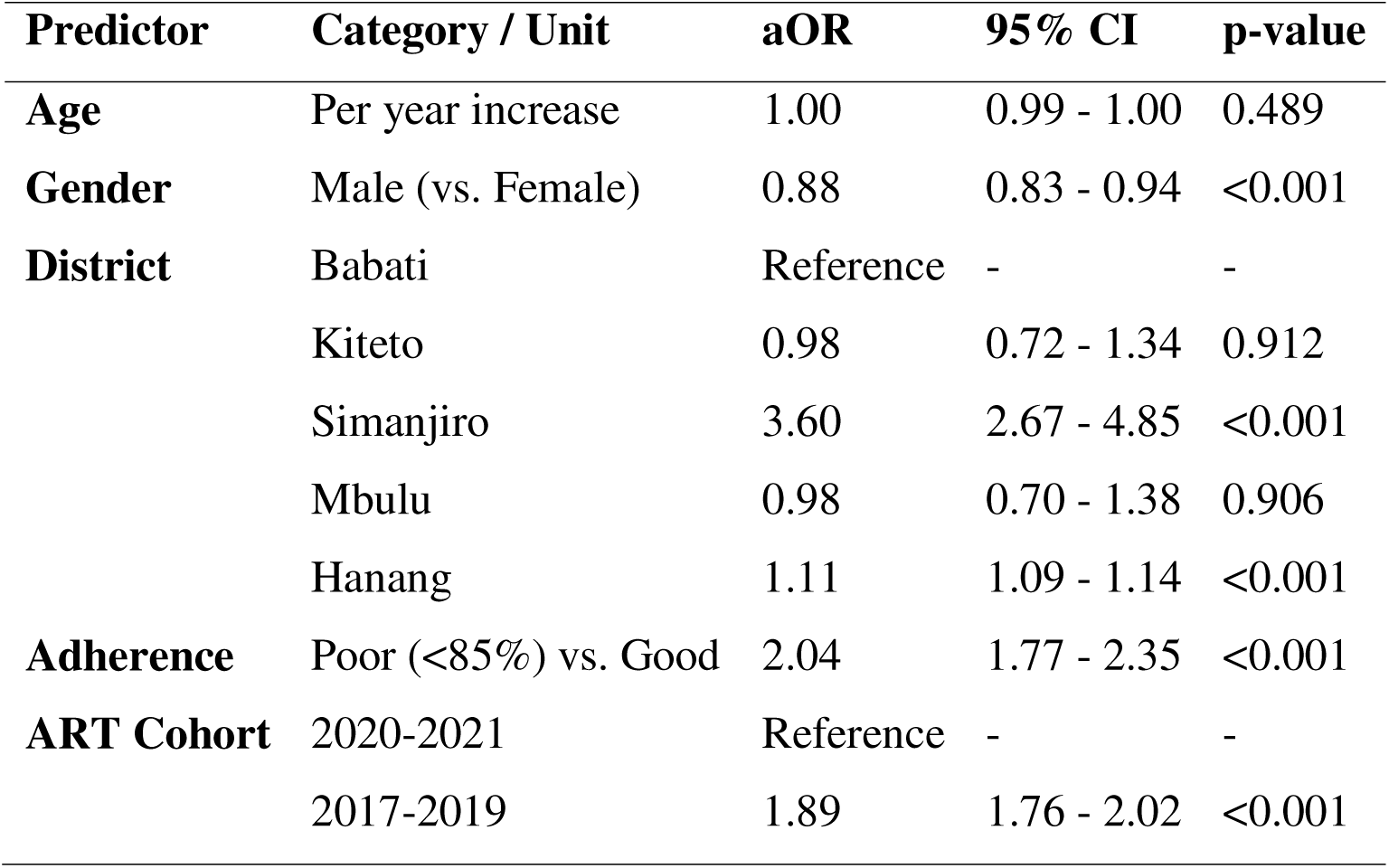
Multivariable Predictors of Loss to Follow-Up (LTFU) Among PLHIV in Manyara region.

## Discussion

This study evaluated pharmacy refill adherence and LTFU among a large cohort of 22,650 PLHIV in the Manyara region of Tanzania. In this setting of relatively low HIV prevalence, we identified critical gaps in retention that appear to be driven by a combination of behavioral adherence patterns and geographic socio-economics.

A central finding of this analysis is the powerful role of pharmacy refill data as a precursor to clinical disengagement. The observed mean adherence of 84% aligns with broader African literature; here, refill levels typically range from 70% to 90% [7,18,19]. However, the finding that poor adherence (<85%) was associated with a more than threefold increase in the odds of LTFU suggests that refill gaps are not merely lapses in medication intake, but are early warning signals of a patient’s impending exit from the formal health system.

Second, district-level differences in adherence and LTFU were substantial. Simanjiro exhibited the lowest adherence and the highest LTFU, whereas Babati and Hanang showed better retention. A sharp spatial heterogeneity across the Manyara region compounds these behavioral risks. The significantly higher odds of LTFU observed in Simanjiro (aOR 3.60) compared to the regional center of Babati suggest that physical and structural barriers are paramount. This geographic risk likely reflects the region’s unique socio-economic landscape, particularly in districts like Simanjiro and Kiteto, which are home to significant nomadic pastoralist populations, such as the Maasai. For these communities, seasonal migration for grazing presents a fundamental structural barrier to accessing fixed-point CTCs [20,21].

Our findings regarding confirmed final status provide a crucial context for this geographic trend. While 32.8% of the cohort met the research definition for LTFU, 23.1% were eventually documented as transfers to other facilities. In nomadic contexts, silent transfers, where patients move to a new area and access a different clinic without formal documentation, are common [20,22]. This suggests that in districts like Simanjiro, a significant portion of the recorded loss may actually represent highly mobile patients maintaining care elsewhere as they follow migratory routes [23]. Such nomadic lifestyles have historically conflicted with traditional healthcare delivery, often resulting in disengagement where patients move beyond the reach of their home facility [22]. Modern systematic evidence corroborates the view that nomadic pastoralists remain among the hardest-to-reach populations in Africa due to a mismatch between migratory routes and the fixed distribution of ART [21]. In Tanzania, healthcare providers continue to face significant challenges in tracking and maintaining follow-up for these mobile clients within the standard national framework [24]. This spatial heterogeneity has been documented in other Tanzanian regions [13,14], suggesting that the NASHCoP should prioritize geographic hotspots for targeted interventions, including community ART refill groups and strengthened cross-district tracing mechanisms.

The timing of ART initiation also emerged as a critical predictor, with patients in earlier cohorts (2017-2019) showing significantly higher attrition than those in more recent cohorts. This trend is consistent with the well-documented phenomenon of treatment fatigue, in which the risk of disengagement accumulates over the course of therapy [12]. Conversely, the improved retention seen in the 2021 cohort likely reflects the successful scale-up of DSD models and multi-month dispensing (MMD) protocols.

Conversely, the improved retention seen in the 2021 cohort likely reflects the successful scale-up of DSD models and multi-month dispensing (MMD) protocols. [15,25]. The marginally lower odds of LTFU among males in our multivariable model contrast with the common literature reporting poorer male retention [15]. This divergence may suggest that regional engagement strategies in Manyara have been particularly effective for men, or that the specific livelihood activities in this region, such as mining or pastoralism, affect men and women differently from more urbanized Tanzanian cohorts.

Collectively, these results support the transition toward more objective, scalable monitoring tools. Unlike self-reported metrics, pharmacy refill data provide an automated evidence base that is easily integrated into the National CTC-2 system [7,9]. By utilizing refill-based algorithms to identify geographic hotspots and individual-level risks, the National HIV program can move toward a more proactive, mobility-friendly model of care. This is particularly vital for nomadic districts, where strategies such as community ART refill groups and cross-district tracing must be prioritized to sustain viral suppression across mobile populations.

## Limitations

Several limitations warrant consideration when interpreting these findings. First, pharmacy refill records measure medication collection rather than actual ingestion, and cannot account for potential diversion or stockpiling. Second, reliance on a single facility network may result in misclassification of loss to follow-up among mobile or nomadic populations who may have obtained refills at facilities outside the study area. This is particularly relevant in geographically vast districts where dispersed settlements and migratory livelihoods create within-district variations in service access that regional mapping cannot fully capture.

Third, the high proportion of missing viral load results, due to intermittent laboratory reporting and programmatic exclusions, limited our ability to correlate refill adherence with biological outcomes. Fourth, the use of routine observational data introduces potential unmeasured confounding from factors such as HIV stigma, mental health, and individual socio-economic status. Finally, as this study was a secondary analysis of electronic records without a qualitative field component, the specific socio-cultural drivers behind the identified geographic hotspots remain to be fully elucidated through future primary research.

## Conclusion

Pharmacy refill adherence and loss to follow-up present significant challenges to the sustainability of HIV treatment programs in the Manyara region. Our findings demonstrate that poor refill adherence is the most potent predictor of clinical disengagement, reinforcing its utility as a high-yield, feasible monitoring metric. Beyond individual behavior, the independent influence of geographic location and ART initiation cohorts highlights persistent structural disparities that traditional facility-based care models have yet to resolve.

The high attrition rate in Simanjiro underscores the vulnerability of nomadic pastoralist populations. These results emphasize the need for the national programme to move beyond fixed-point clinics. By integrating automated refill metrics into national systems and expanding community-based delivery models that accommodate migratory patterns, the program can better support mobile populations. Such data-driven, localized interventions are critical for closing the retention gap and achieving the national 95-95-95 targets.

## Declarations

### Ethics approval and consent to participate

This study used de-identified routine program data from the NASHCoP. Ethical approval for the secondary analysis of CTC-2 data was obtained from the Muhimbili University of Health and Allied Sciences (MUHAS) Institutional Review Board under protocol number DA.282/298/01L/923 issued in February 2024.

### Consent for publication

Not applicable. The manuscript does not include any identifiable personal data.

### Availability of data and materials

The NASHCoP owns the dataset supporting the findings of this study and contains sensitive clinical information that cannot be publicly shared. Aggregated or de-identified data may be made available from the corresponding author upon reasonable request and with permission from the NASHCoP.

### Authors’ contributions

MBK led the data preparation, statistical analysis, and drafting of the initial results.

RZS conceived the study, guided the analytical framework, interpreted the findings and finalized the manuscript.

JM contributed to the study’s conceptualization and early methodological design. GRS provided technical support for data visualization and analytical tools.

VS and PN facilitated access to programmatic data and provided oversight of the NASHCoP. All authors reviewed and approved the final manuscript.

## Acknowledgements

We acknowledge the National AIDS and Sexually Transmitted Infections Control Programme (NASHCoP) for granting access to the routine CTC-2 data used in this study. We also thank the regional and district HIV coordinators in Manyara for their support in validating programmatic indicators.

## Disclaimer

The findings and conclusions presented in this article are those of the authors and do not necessarily represent the official views of the NASHCoP, the Ministry of Health, or any affiliated institutions.

